# DNA methylomes derived from alveolar macrophages display distinct patterns in latent tuberculosis - implication for interferon gamma release assay status determination

**DOI:** 10.1101/2021.03.16.21253725

**Authors:** Isabelle Pehrson, Jyotirmoy Das, Nina Idh, Lovisa Karlsson, Helena Rylander, Hilma Hård af Segerstad, Elsa Reuterswärd, Emma Marttala, Jakob Paues, Melissa Méndez-Aranda, César Ugarte-Gil, Maria Lerm

## Abstract

Host innate immune cells, including alveolar macrophages, have been identified as key players in the early eradication of *Mycobacterium tuberculosis* and in the maintenance of an anti-mycobacterial immune memory, which is believed to be induced through epigenetic changes. The aim of the study was to elucidate whether exposure to *M. tuberculosis* induced a different DNA methylation pattern of alveolar macrophages and pulmonary T lymphocytes.

Alveolar macrophages and T lymphocytes were isolated from induced sputum obtained from individuals living in Lima, which is an area high endemic for tuberculosis. To determine the latent tuberculosis infection status of the subjects, an interferon-γ release assay was performed. We evaluated the DNA methylomes of the alveolar macrophages and T lymphocytes using the Illumina Infinium Human Methylation 450K Bead Chip array, revealing a distinct DNA methylation pattern in alveolar macrophages allowing the discrimination of asymptomatic individuals with latent tuberculosis infection from non-infected individuals. Pathway analysis revealed that cell signalling of inflammation and chemokines in alveolar macrophages play a role in latent tuberculosis infection. In conclusion, we demonstrated that DNA methylation in alveolar macrophages can be used to determine the tuberculosis infection status of individuals in a high endemic setting.

## Introduction

Infection with *Mycobacterium tuberculosis*, the causative agent of tuberculosis (TB), can result in active disease or latent infection. Globally, two billion individuals are currently estimated to have a latent TB infection (LTBI), and 5-15% of those will develop active TB during their lifetime ^1-4^. In the rest of TB-exposed individuals, the innate immune system will eliminate the bacteria before the onset of an adaptive immune response, with no detectable evidence of a persistent immune response, referred to as early clearance ^5,6^.

*M. tuberculosis* is primarily transmitted via airborne droplets and inhaled into the airways where it establishes an infection that could develop into pulmonary TB ^1^. When *M. tuberculosis* reaches the alveoli within the lungs, the pathogen is phagocytosed by alveolar macrophages, a subgroup of lung tissue-resident macrophages, which form part of the innate immune system and are among the first host cells that the bacteria encounter ^7,8^. Subsequently, lung T lymphocytes are recruited and begin to engage in immune responses against the pathogen ^8,9^.

The only TB-vaccine available today is the *M. bovis* Bacillus Calmette-Guèrin (BCG). The vaccine is effective in protecting infants and small children against miliary TB and meningitis but is unsuccessful in protecting adults against pulmonary TB ^10,11^. In addition to its protective effect against TB in children, the vaccine also protects them against unrelated pathogens and even reduce general mortality in children, which could be explained by epigenetic reprogramming of innate immune cells, referred to as trained immunity ^12-16^. The trained innate immune system is in part regulated by DNA methylation, an epigenetic modifier which regulates various processes within the cell, including anti-inflammatory responses ^17,18^.

For the diagnosis of LTBI, the best options today are the tuberculin skin test (TST) and interferon-γ (IFN-γ) release assay (IGRA) ^1,19^. The IGRAs detect IFN-γ secondary to T lymphocyte activation and secretion as a response to exposure to *M. tuberculosis*-specific proteins. The IGRA T-SPOT.*TB* has shown a high reproducibility in serial testing and to have higher sensitivity than other available tests ^20,21^.

LTBI is a dynamic state that can progress to active tuberculosis under immunesenescence or immunosuppression, since it reflects a balance between host immune responses and the bacilli. Thus, LTBI can be viewed as a spectrum, with variable levels of bacterial control and risk of progression to active TB infection ^22-24^. Even though attempts have been made to identify biomarkers of LTBI and LTBI progression, including DNA methylation events in peripheral immune cells of active and latent TB patients ^25,26^, no study to our knowledge has investigated DNA methylation in cells of the lung compartment of IGRA-positive latently infected individuals.

Biosignatures of TB have been proposed to lay ground for next generation of diagnostics for TB ^27,28^ and accumulating literature have been published studying possible TB biosignatures, based on e.g., global methylation analysis, transcriptional profiling and RNA sequencing enrichment ^29-32^. However, no biomarker or signature of active or latent TB has been successful when generalized or applied in larger sample sizes and biosignature based diagnostics tools are still unavailable in the clinic.

In this study, we isolated alveolar macrophages from individuals residing in a Lima, Peru which has a high prevalence of tuberculosis. We performed DNA methylation analyses to identify changes in the methylome and detected 265 significant differently methylated CpG sites (DMCs) when comparing individuals with LTBI with IGRA-negative non-TB (NTB) individuals. Further analysis of the DMCs revealed their importance in several pathways of the innate immune system.

## Methods

### Ethical approval

Ethical approval was obtained from the Institutional Committee on Research Ethics “Comité Institucional de Ética en Investigación” (CIEI) of the Cayetano Heredia National Hospital, N^o^ 082-018 for the inclusion of patients and Universidad Peruana Cayetano Heredia Institutional Review Board N° 101773 for the inclusion of healthy subjects.

### Subjects

Peru has a high prevalence of TB and a heavy burden of multidrug-resistant (MDR) and extensively drug-resistant (XDR) TB ^33,34^. With reference to the increased risk of Peruvian health care workers and medical students to acquire a TB infection ^35-38^, subjects were recruited from the from the Universidad Peruana Cayetano Heredia (UPCH) and the Hospital Nacional Cayetano Heredia in Lima, Peru during April, October and November 2018. 26 individuals aged 18-40 years were included in the study following oral and written consent. Based on the IGRA results, the subjects were divided into IGRA-positives (LTBI group) and IGRA-negatives (here defined as NTB group). DNA from alveolar macrophages and T lymphocytes, obtained via sputum induction, were isolated from both groups and the number of differentially methylated CpG-sites (DMCs) assessed. For the alveolar macrophages, pathway analysis and cluster analysis were performed. An auxiliary heatmap analysis was performed with the addition of two drug resistant tuberculosis (DRTB) patients, included in June 2019. Following cell isolation and DNA extraction, one HLA-DR and one CD3 sample from two different subjects (both NTB) had to be excluded due to insufficient DNA amount.

### IF N-γ release assay

The latent TB infection status of all subjects except for the active TB patients was evaluated with an IGRA. The TB infection status of the active TB patients was based on current medical records. Following venepuncture, the IGRA (T-SPOT.*TB*, Oxford Immunotec) was performed, measuring the IFN-γ release of the peripheral immune cells, per the manufacturer’s protocol. The T-SPOT.*TB* result is based on the number of spot-forming cells (memory T cells producing IFN-γ, sensitized in response to the mycobacterial antigens ESAT-6 and CFP-10).

### Sputum induction and processing

Sputum specimen were collected and processed as previously described ^39^, with some modifications. The lung immune cells were obtained from each subject (here, salbutamol was avoided) after the nebulization of a hypertonic saline solution (4%) three times, provoking a profound cough via osmosis, producing pulmonal secretions. The coughing was preceded by an oral cleaning procedure. After the collection of the expectorate via sputum induction, the cell-rich samples were transferred to various cell-culture dishes and accumulations of mucus of immune cells, referred to as “plugs”, were transferred to tubes using forceps. Following the plug collection, the volume of the sample was estimated and a solution containing phosphate buffer solution (PBS) and 0.1% dithiothreitol (DTT) was added to the sample equal to four times the volume of the sample, to provide a reducing agent to decrease sample viscosity. Following this, the sample was diluted with PBS, and rocked for 5 minutes before filtrated through a 50 µm cell strainer into a new tube to remove contaminating squamous cells and centrifuged for 5 minutes.

### DNA extraction of isolated alveolar macrophages and T lymphocytes

Sikkeland *et al*. have previously described the method for the isolation of HLA-DR positive alveolar macrophages ^39,40^. The alveolar macrophages and T lymphocytes (CD3 positive cells) were isolated according to our protocol ^39^ using superparamagnetic beads coupled with anti-human CD3 and Pan Mouse IgG antibodies and HLA-DR/human MHC class II antibodies (Invitrogen Dynabeads). An initial positive selection was done with CD3 beads followed by a positive HLA-DR selection. Bead coating and cell isolation was performed according to manufacturer’s protocol. The DNA and RNA were extracted from the lung immune cells using the AllPrep DNA/RNA Mini Kit (Qiagen), per the manufacturer’s instructions, and quantified using the spectrophotometer NanoDrop 2000/2000c (Thermo Scientific).

### DNA methylation data analysis

Genome-wide DNA methylation analysis of the DNA from the two cell types (HLA-DR and CD3 positive cells) was performed using the Infinium Human Methylation 450K Bead Chip array (Illumina) as per the manufacturer’s instructions. The technique is based on quantitative genotyping of C/T polymorphism which is generated by DNA bisulfite conversion and allows for the assessment of over 450 000 methylation sites within the whole genome. DNA was converted and amplified and subsequently fragmented and hybridized to the Infinium HumanMethylation450 Bead Chip. The procedure was conducted at the Karolinska Institute, Sweden.

### Computational analysis

The IDAT files (RAW files that contain green and red signals from methylated and unmethylated CpG sites) were analyzed using the R programming language (v3.6.3) (R Foundation for Statistical Computing: Vienna, Austria) and Chip Analysis Methylation Pipeline (*ChAMP*) analysis package ^41^. The following filters were applied: i) detection of *p*-value (<0.01), ii) remove non-CpG probes, iii) SNP removal, iv) remove multi-hit probes, v) remove all probes in X and Y chromosomes ^41^. After filtering, quality control was performed, and normalization of the data was made using the BMIQ method. The β-values and M-values of the samples were calculated against each probe per sample. Differential methylation analysis of CpG sites (DMCs) between the sample groups was then performed using Bonferroni-Hochberg (BH)-corrected *p*-value (*p*-value_BH_) < 0.05. The DMCs were annotated with the Illumina manifest file using GRCh37/hg19 database.

The statistically significant (*p*-value_BH_ < 0.05) was used to create the static volcano plot with the |log2FC| ≥0.3 using the *EnhancedVolcano plot* package package ^42^ in R. The hierarchical cluster analyses were performed from the β-values of the significant DMCs from each participant using the *hclust* function of the *ade4/ape* package ^43^ in R. The R packages *ComplexHeatmap* ^44^ was used to create the heatmap from individual β-values of DMCs (*p*-value_BH_ < 0.05 and log_2_ fold change ≥ |0.4|). For the CD3 positive cells we found 0 DMCs (*p*-value_BH_ < 0.05) (not shown) and thus no other analyses were performed on the CD3 positive cells. The design matrix of the DNA methylation analysis was “LTBI - NTB” meaning that a positive log_2_ fold change value equals hypermethylated DMC in the LTBI group. Negative log_2_ fold change value means NTB DMCs are hypermethylated. Pathway enrichment analysis was performed using Signaling Pathway Impact Analysis (SPIA) ^45^ enrichment analysis and *pathfindR* ^46^, based on information from the Kyoto Encyklopedia of Genes and Genomes (KEGG) ^47^. Via the SPIA enrichment analysis, based on pathway information from KEGG, we used the expression rates of the LTBI vs NTB DMGs to calculate signaling pathway topology. The R package *pathfindR* is a tool for pathway enrichment analyses that uses active subnetwork to calculate the pathway enrichment. Using the enriched DMGs separating the LTBI from NTB, we identified active subnetworks and performed pathway enrichment analyses on the identified subnetworks. Via a scoring function, the overall activity of each pathway in each sample was estimated.

## Results

### Subject characteristics

In order to identify possible DMCs to reflect TB-exposure, we included healthy subjects with presumable high and low exposure to TB residing in a high endemic area in Lima, Peru and subsequently divided into two groups based on their IGRA result (Fig. 1). Among the healthy subjects that were included, six we identified as IGRA-positive (LTBI) and 19 as IGRA-negative individuals (NTB). Demographics, body metrics, Bacillus Calmette-Guèrin (BCG)- and smoking status are summarized in Table 1. The patients (DRTB) were hospitalized and were suffering from MDR-TB and XDR-TB, respectively (Table 2). One patient was smear microscopy positive and suffered from cough, dyspnea, and night sweats while the other was asymptomatic and smear microscopy negative at the time. One patient had received the MDR-antibiotic regimen since 2016. The other patient had a TB disease history of over 10 years, with susceptible TB since 2008 and a later progression to MDR-TB (2016) and XDR-TB (2019). Both patients were treated with multiple antibiotics at the time of inclusion.

**Figure 1.**
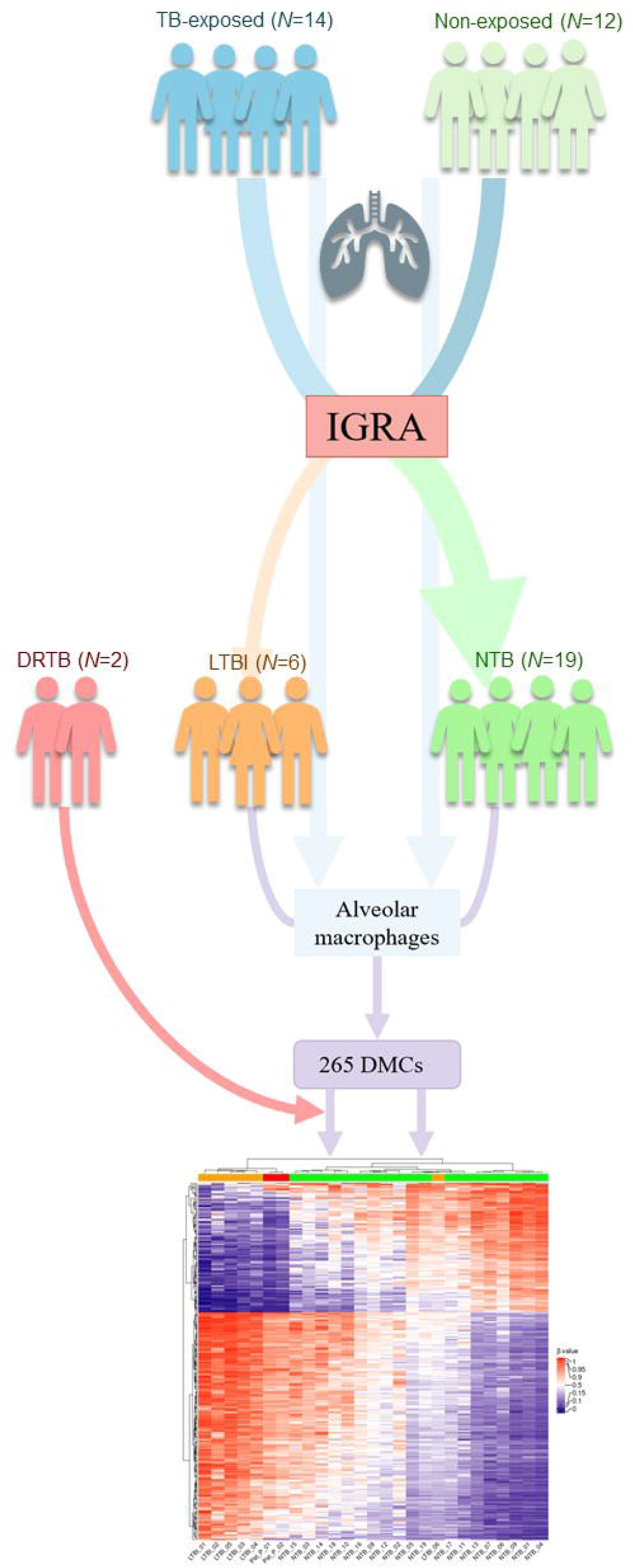
A flow chart of the study design. TB = tuberculosis. IGRA = interferon-γ release assay. LTBI = latent tuberculosis infection. NTB = non-tuberculosis. DRTB = drug resistant tuberculosis. DMCs = differentially methylated CpG-sites.

**Table 1.** Demographic and clinical data of the LTBI and NTB study population.

|  | **LTBI**<br>**(N=6)** | **NTB**<br>**(N=19)** | **Total**<br>**(N=25)** |
| --- | --- | --- | --- |
| Age (mean) | 23 (19-26) | 25 (18-40) | 25 (18-40) |
| Sex (male/female) | 5/1 | 10/9 | 15/10 |
| Weight (kg) | 78.2<br>(54-94) | 63.2<br>(47-85) | 66.8<br>(47-94) |
| Height (cm) | 175.7<br>(168-183) | 165.4<br>(148-178) | 167.8<br>(148-183) |
| BMI (kg/m<sup>2</sup>) | 25.2<br>(19.1-29.3) | 22.9<br>(19.1-28.7) | 23.5<br>(19.1-29.3) |
| Smoker/non-smoker | 2/4 | 1/18 | 3/22 |
| IGRA result<br>(positive/negative) | 6/0 | 0/19 | 6/19 |
| History of TST-positivity<br>(yes/no/ND) | 1/4/1 | 3/6/10 | 4/10/11 |
| BCG (vaccinated/non-vaccinated) | 6/0 | 16/3 | 22/3 |
| Included as TB-exposed/non-exposed | 5/1 | 9/10 | 14/11 |
| Previous TB-treatment<br>(yes/no) | 0/6 | 1/18 | 1/24 |
| Previous dust exposure<br>(yes/no) | 0/6 | 0/19 | 0/25 |

LTBI = latent tuberculosis infection. NTB = non-tuberculosis. BMI = body mass index. BCG = Bacillus Calmette-Guérin. ND = no data. TB = tuberculosis. TST = tuberculin skin test.

**Table 2.** Demographic and clinical data of the two drug resistant pulmonary tuberculosis patients.

|  | **DRTB (N=2)** |
| --- | --- |
| Age (mean) | 30 (28-32) |
| Sex (male/female) | 2/0 |
| Weight (kg) | 61.5 (58-65) |
| Height (cm) | 173 (166-180) |
| BMI (kg/m<sup>2</sup>) | 20.6 (20.1-21.1) |
| Smoker/previous smoker/non-smoker | 0/1/1 |
| BCG (vaccinated/non-vaccinated/ND) | 1/0/1 |
| Previous TB-treatment (yes/no) | 2/0 |
| Previous dust exposure (yes/no) | 1/1 |
| MDR-TB/XDR-TB | 1/1 |
| Smear microscopy (positive/negative) | 1/1 |
| Years of TB-disease | 7 (3-11) |
| Duration of MDR-TB-treatment (years) | 4.5 (3-6) |

DRTB = drug-resistant TB. BMI = body mass index. BCG = Bacillus Calmette-Guérin. ND = no data. TB = tuberculosis. MDR = multi-drug resistant. XDR = extensively drug resistant.

### Differential DNA methylation analysis identified DMCs to distinguish LTBI from NTB in alveolar macrophages

To determine whether LTBI and NTB can be distinguished based on the methylation pattern, we compared the alveolar macrophage and T lymphocyte DNA methylome data obtained from LTBI and NTB subjects. The analysis revealed 265 DMCs with a *p*-value_BH_ < 0.05 and log_2_ fold change value ≥|0.3| for alveolar macrophages (not shown). The DMCs differentiating LTBI from NTB are presented as DMCs with their equivalent DMGs in a volcano plot (Fig. 2).

**Figure 2.**
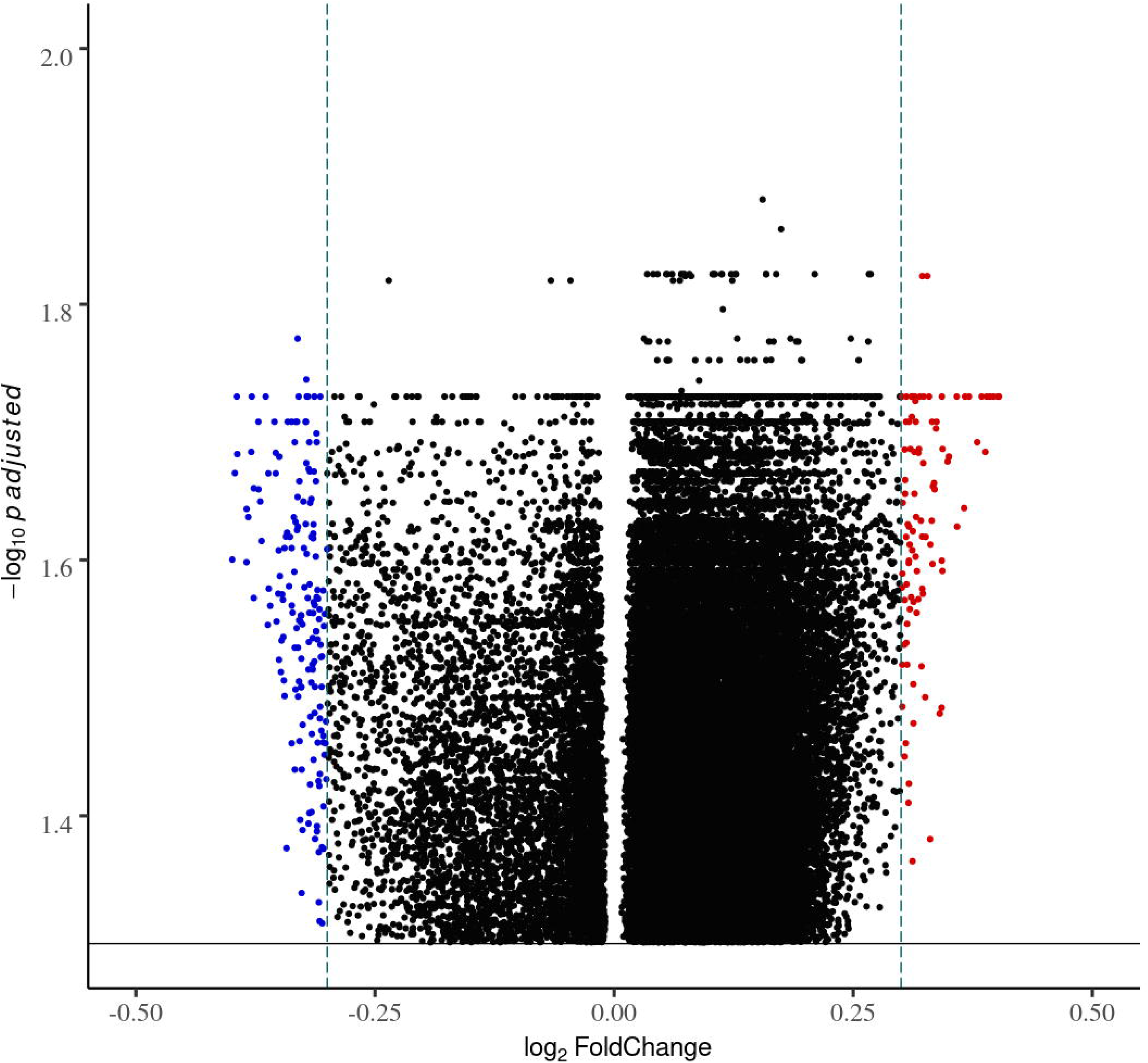
Volcano plot of the differentially methylated CpG-sited (DMCs) separating the latent tuberculosis infection (LTBI) from the non-tuberculosis (NTB) group. The plot is showing the distribution of the DMCs fold changes in LTBI relative to NBT in alveolar macrophages. Each dot represents one CpG site (*p*-value_BH_ < 0.05). x-scale = log_2_FC, y-scale = -log_10_ p adjusted; blue CpGs = hypomethylated; red CpGs = hypermethylated; black CpGs = significant but |log_2_FC|<0.3 horizontal black line in parallel to x-axis = *p*-value_BH_ < 0.05, and vertical green lines parallel to y-axis = |log_2_FC|≥0.3.

In total, 36 055 DMCs was found to distinguish LTBI from NTB without any log_2_ fold change value cut-off (not shown). The same DNA methylation analysis performed with the lung T lymphocytes resulted in 0 DMCs (*p*-value_BH_ < 0.05) (not shown). Thus, no further analysis was performed on the T lymphocytes. No DMCs were found between the subjects included as TB-exposed and non-exposed (not shown).

### A heatmap analysis revealed a distinct DNA methylation pattern of LTBI and drug resistant TB in alveolar macrophages

After demonstrating that the LTBI and NTB groups had distinct DNA methylation traits, knowing that heterogenicity of the epigenome of LTBI subjects has previously been demonstrated ^48^ and that LTBI can be viewed as a spectrum ^22^, we performed a heatmap analysis based on the top DMCs of the DNA methylation analysis (265 DMCs, *p*-value_BH_ <0.05, log_2_ fold change ≥|0.3|) that showed that the LTBI group, with the exception of one participant, clustered into one group (Fig. 3a). In order to investigate whether we could identify any TB-like subjects within the diverse LTBI group, as previously demonstrated by Estévez *et al*. ^48^, we recruited two individuals with TB and with ongoing treatment to the study (Table 2). The cluster analysis revealed that the DRTB group clustered with the 5 LTBI subjects (Fig. 3a). A subsequent principal component analysis (PCA, Fig. 3b) revealed that the DRTB, despite the similarities with the LTBI group in the cluster analysis, formed a separate population, demonstrating that the three conditions are clearly separated by the identified DNA methylation signature.

**Figure 3a.**
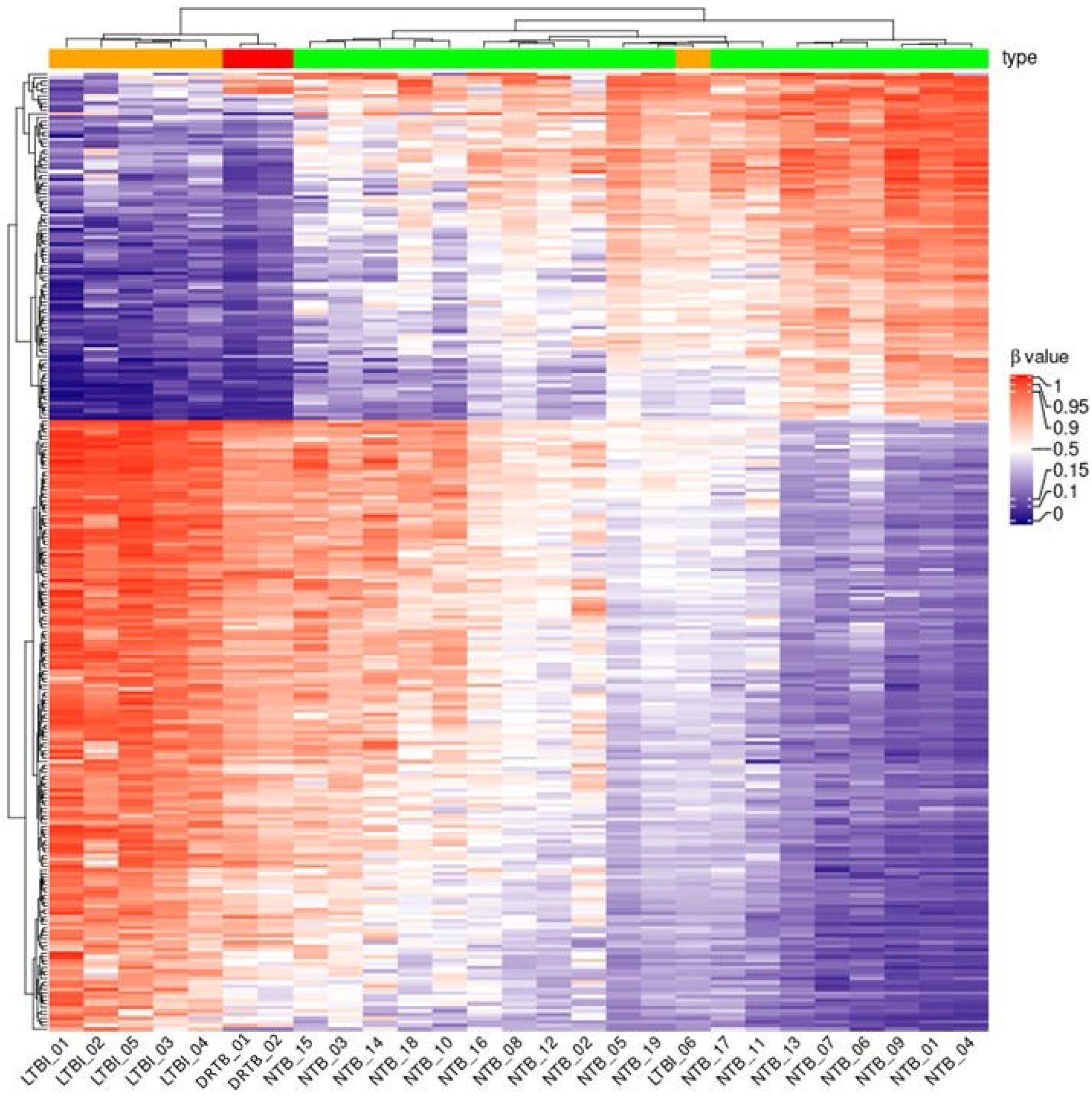
Heatmap of the LTBI, NTB and DRTB groups. The heatmap is plotted from the β-values of the top 265 CpGs (log_2_ fold change ≥|0.3|, *p*-value_BH_ < 0.05) differentiating the LTBI and NTB groups. The drug-resistant TB patients (DR-TB) are added to the analysis. The color scale represent the β-value (from 0 (blue) to 1 (red)) of each CpG site. Orange: LTBI = latent tuberculosis infection, green: NTB = non-tuberculosis, red: DRTB = drug-resistant tuberculosis as shown in the bar plot of the heatmap. Cluster dendrogram is calculated using the Euclidean distance method.

**Figure 3b.**
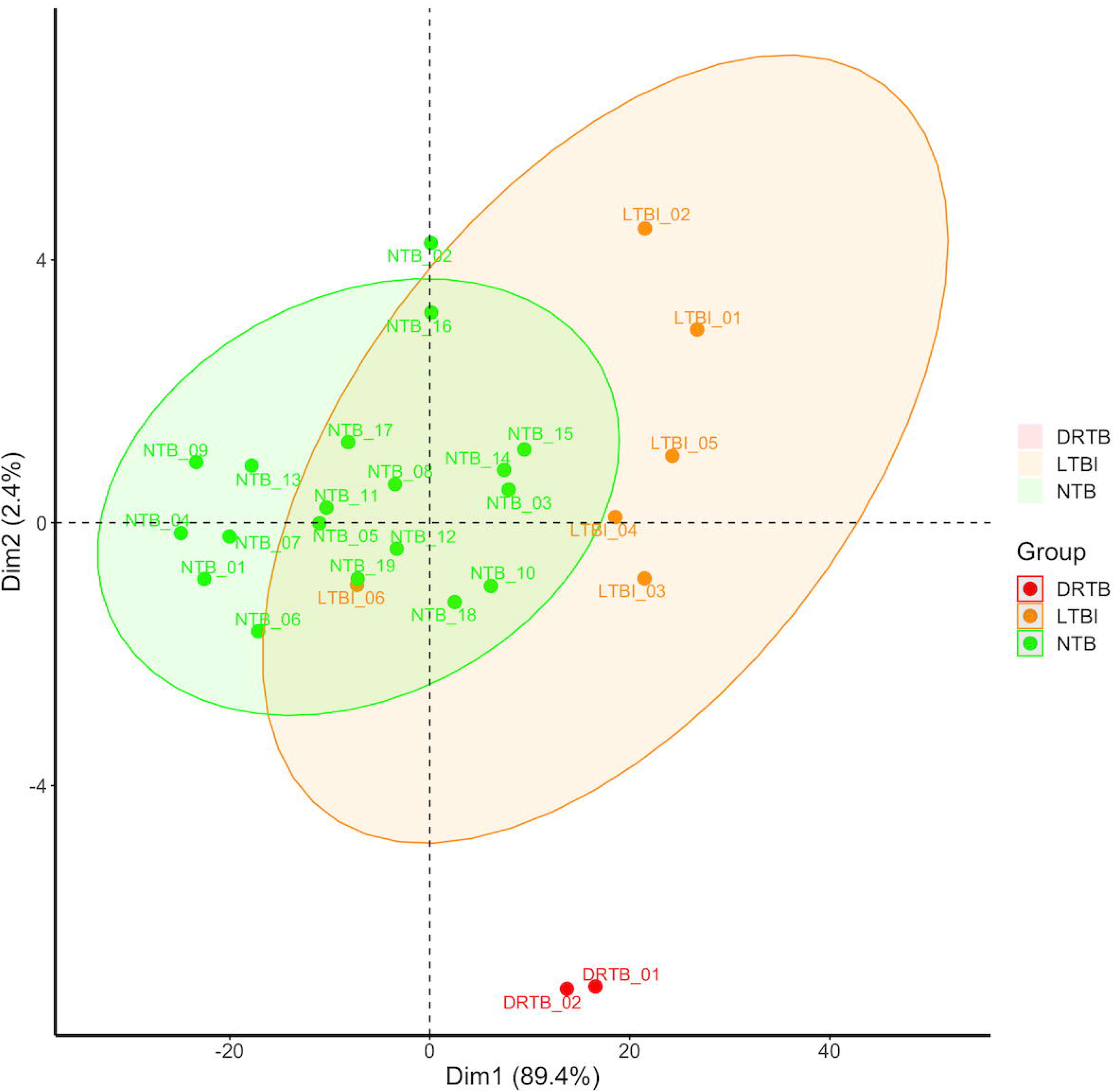
Principal components analysis (PCA) of the LTBI, NTB and DRTB group. The PCA plot is generated from the β-values of the top 265 CpGs (log_2_ fold change ≥|0.3|, *p*-value_BH_ < 0.05) differentiating the LTBI and NTB groups, with the addition of the β-values of the DRTB patients of those DMCs.

### Biological processes involved in latent tuberculosis infection

Next, we investigated the biological processes involved in LTBI, based on the genes found in the differential DNA methylation analysis. We performed pathway enrichment analyses of the alveolar macrophages to identify the common pathways that distinguished the LTBI samples from the NTB and found pathways involved in the spliceosome, Wnt signaling, mTOR signaling, regulation of actin cytoskeleton, ribosome, extracellular matrix organization, MAPK signaling, cellular senescence, TGF-β signaling and the phosphatidylinositol signaling system (Fig. 4).

**Figure 4.**
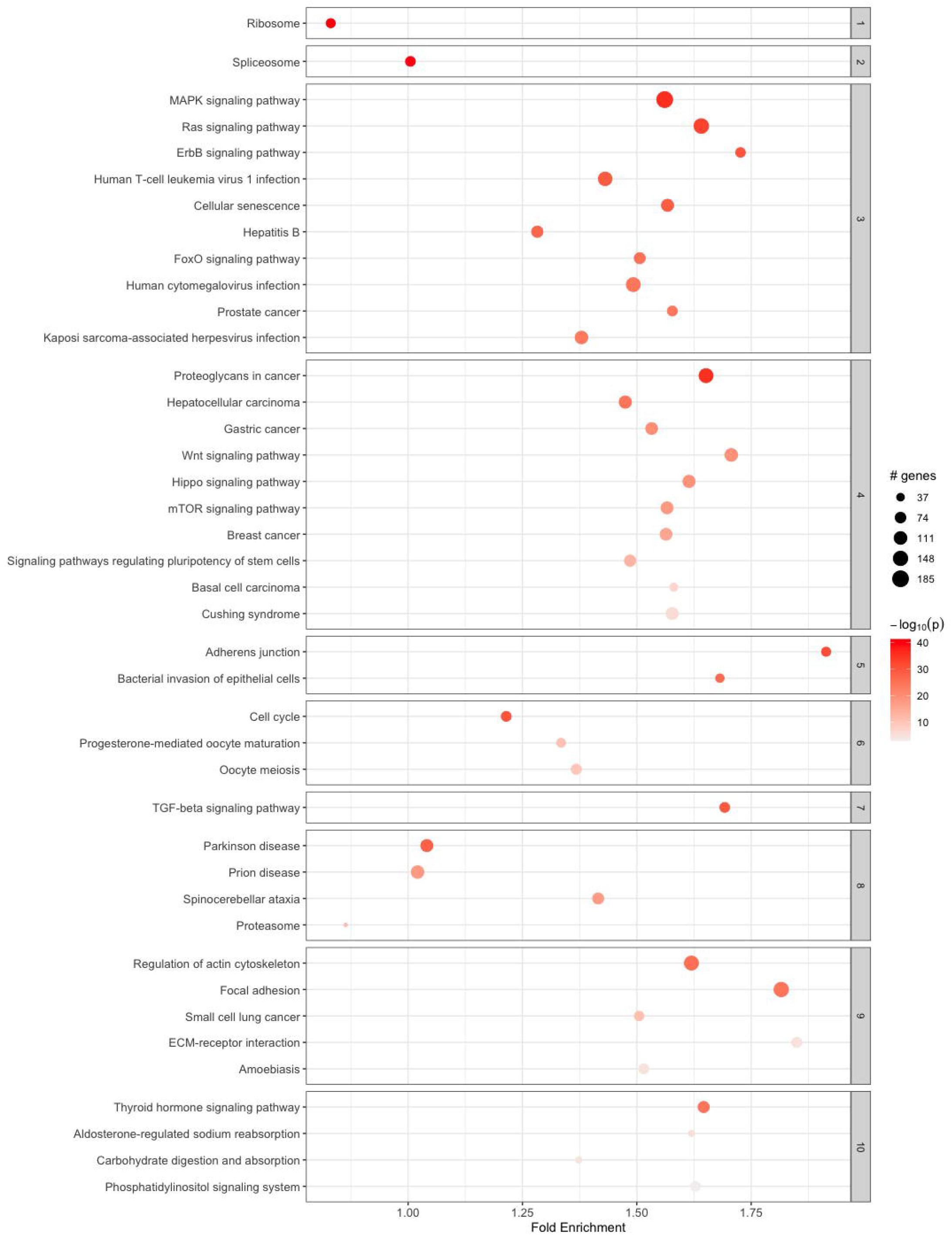
Top 10 clusters of the enriched pathways from pathfindR based on the differentially methylated genes (DMGs) differentiating LTBI from NTB. The dot plot is demonstrating the most relevant pathways for the alveolar macrophages when comparing the LTBI with the NTB group. The gene count is presented as circles, increasing with elevating number of gene count. The significance level is presented as a color scale (-log_10_(*p*)).

In addition, SPIA pathway enrichment analysis was performed, corroborating the enrichment of innate immune-related pathways, including chemokine signaling pathway, antigen processing and presentation, *Staphylococcus aureus* infection, the NOD-like receptor signaling pathway and intestinal immune network for IgA production (Table S1).

### Counter DNA methylation status of genes persistently hypomethylated in BCG responders

In our previous study, PBMCs obtained from BCG-vaccinated individuals, whose macrophages displayed an enhanced anti-mycobacterial response, displayed persistent hypomethylation in six genes, among them the gene encoding IFN-γ ^49^. In order to estimate whether these genes’ DNA methylation status of the BCG study was coherent with the LTBI data in the present study, we compared the β values of these genes in the different studies. Contrary to the observations in the BCG vaccinees, the six genes were found to be hypermethylated in LTBI (Table S2).

## Discussion

In this study, we found that five out of six individuals diagnosed with LTBI based on a positive IGRA displayed a distinct DNA methylation pattern of alveolar macrophages. The reason why one of the LTBI individual’s signature aligned with the NTB group remains obscure, but it is tempting to speculate that this individual had cleared a previous latent TB infection, allowing the LTBI-specific DNA methylome signature to fade while the adaptive immune memory against TB antigens endured. However, further studies are needed to provide evidence for such a relationship. A limitation with the present study is that no information on previous IGRA test results was available, and future studies will dissect the relationship between the timing of IGRA conversion and the intensity of the DNA methylome signature.

In our previous study on DNA methylation alterations in response to the BCG vaccine, we identified CpG sites in six gene promoters, one of which belonging to the *IFNG* gene (encoding IFN-γ), as being persistently more accessible for transcription (hypomethylated) in BCG responders. In contrast, the present study showed that the very same six CpG sites were hypermethylated in the LTBI group ^49^. We hypothesize that the transcription accessibility of these six genes has importance for the clearance of *M. tuberculosis*, reflective of a beneficial BCG response ^49^. LTBI is a manifestation of lost mycobacterial control by innate mechanisms and possibly a consequence of hypermethylation of for example IFN-γ, which has a non-disputable importance for the control of TB ^50^.

While being a risk factor for developing active TB, LTBI has been shown to convey protection against *M. tuberculosis* reinfection, partly explained by heterologous immunity and the concomitant activation of the innate immune system ^51,52^. The protective mechanisms of LTBI were associated with activation of alveolar macrophages and accelerated recruitment of *M. tuberculosis*-specific T cells to the lung and enhanced cytokines production ^52^, compatible with our results where we showed an overrepresentation of pathways involved in TGF-β, NOD-like receptor (NLR) and chemokine signaling. The activation of the NLRP3 inflammasome, activated via *NLRP3* promotor demethylation, has previously been identified as an important regulatory function in TB ^53^. Interestingly, *NLRP3* was differentially methylated between the LTBI and NTB at 3 CpG sites in our data and we identified NLR signaling as an activated pathway in LTBI. Increased NLPR3 inflammasome activation enhances the anti-mycobacterial capacity of macrophages ^54^ and prevents development of active pulmonary TB ^55^.

Methylation of the *VDR* promoter (regulating Vitamin D receptor expression) has previously been demonstrated to be of importance in TB ^56-58^, especially for disease progression and risk. Vitamin D enhances macrophage capacity to control intracellular *M. tuberculosis* ^59,60^. In our data, we found three differentially methylated *VDR* CpG sites, all hypermethylated in the LTBI group, pointing towards downregulation of VDR responsiveness in alveolar macrophages from subjects with LTBI, which may impair their ability to kill intracellular mycobacteria ^61^.

Early clearance is observed in about 25% of individuals who are living with considerable exposure to TB ^62-64^, and epigenetic mechanisms related to trained immunity could contribute to the elimination of the infection without any need to trigger an adaptive immune response ^65^. We propose that the LTBI group has been unsuccessful in clearing the bacteria and that the epigenetic differences we identify between the LTBI and NTB group.

The heatmap analysis revealed that the DNA methylation pattern of alveolar macrophages of DRTB patients, who were on MDR treatment at the timepoint of inclusion, displayed similar pattern with the LTBI individuals, however they formed a distinct cluster the PCA analysis. Since the DRTB patients had been on TB treatment for an extended period of time, our study does not reveal the DNA methylome signature of active pulmonary TB, but suggests that DNA methylome analysis is a promising approach to study the spectrum of TB. Therefore, longitudinal studies are needed that cover DNA methylation changes of alveolar macrophages before the time point of IGRA conversion over the development of active TB and the TB treatment until cure. The results will allow dissection of the unique DNA methylation signatures for each state of TB and could be further developed as tools for clinical TB diagnosis, decision-making in LTBI infected subjects and TB treatment monitoring.

### Data and materials availability

Sequencing datasets will be made publicly available upon acceptance and prior to final publication.

## Data Availability

Sequencing datasets will be made publicly available upon acceptance and prior to final publication.

## Acknowledgements

We owe our thanks to the Bioinformatics and Expression Analysis core facility (BEA), which is supported by the board of research at the Karolinska Institute and the research committee at the Karolinska hospital. The study was funded through a grant from the Swedish Research Council N^o^ 2018-04246 and the Consejo Nacional de Ciencia, Tecnología e Innovación Tecnológica CONCYTEC and Cienciactiva (N° 106-2018-FONDECYT). The computations and data handling were enabled by resources provided by the Swedish National Infrastructure for Computing (SNIC) at National Supercomputing Centre (NSC), Linköping University partially funded by the Swedish Research Council through grant agreement N^o^ 2018-05973. We also owe thanks to the support by grants from the World Infection Fund. J.D is a postdoctoral fellow supported through the Medical Infection and Inflammation Center (MIIC) at Linköping University. We direct our gratitude to all the subjects for donating samples. We are also grateful for the help of Ronald Cadillo Hernandez and Rodrigo Cachay Figueroa.

## Authors Contribution

M.L, N.I, J.P, C.U-G and M.M-A wrote the ethical application and conceived the study. I.P, E.M, H.R, E.R and H.H performed all sample collections. J.D and L.K preformed the bioinformatic analyses. I.P, M.L, J.D and L.K wrote the manuscript. M.M-A and C.U-G supervised the project in Peru.

## Consent for publication

All authors have approved to the final version of the manuscript.

## Table legends

**Supplementary table S1.** Table of 7 significantly over-represented pathways identified via the generation with the Signaling Pathway Impact Analysis (SPIA) enrichment analysis. The pathway information is based on Kyoto Encyklopedia of Genes and Genomes (KEGG) generated pathways. Global *p*-value = P_G_. FDR = false discovery rate.

**Supplementary table S2.** Comparison of the DNA methylation status of 6 genes between our data and data from Verma *et al*. (2017) ^49^.

