## Supplementary table S1 for "DNA methylomes derived from alveolar macrophages display distinct patterns in latent tuberculosis - implication for interferon gamma release assay status determination"

**Supplementary table S1**. Table of 7 significantly over-represented pathways identified via the generation with the Signaling Pathway Impact Analysis (SPIA) enrichment analysis. The pathway information is based on Kyoto Encyklopedia of Genes and Genomes (KEGG) generated pathways. Global *p*-vale = P_G_. FDR = false discovery rate.

| **Name** | **KEGG ID** | **P_Size_** | **NDE** | **P_G_** | **P_G_FDR** | **Status** |
| --- | --- | --- | --- | --- | --- | --- |
| Chemokine signaling pathway | 4062 | 110 | 110 | 6.60E-05 | 0.003015387 | Inhibited |
| Antigen processing and presentation | 4612 | 39 | 39 | 6.60E-05 | 0.003015387 | Inhibited |
| Systemic lupus erythematosus | 5322 | 32 | 32 | 6.60E-05 | 0.003015387 | Inhibited |
| Olfactory transduction | 4740 | 104 | 104 | 0.007907755 | 0.270840618 | Activated |
| Staphylococcus aureus infection | 5150 | 30 | 30 | 0.014429216 | 0.395360524 | Inhibited |
| NOD-like receptor signaling pathway | 4621 | 26 | 26 | 0.04662651 | 0.912547408 | Activated |
| Intestinal immune network for IgA production | 4672 | 22 | 22 | 0.04662651 | 0.912547408 | Inhibited |
