## Supplementary table S2 for "DNA methylomes derived from alveolar macrophages display distinct patterns in latent tuberculosis - implication for interferon gamma release assay status determination"

**Supplementary table S2**. Comparison of the DNA methylation status of 6 genes between our data and data from Verma *et al*. (2017) ^49^.

| **Gene symbol** | **Description** | **Methylation status in our data** | **Methylation status in in responders Verma *et al.* (2017)** |
| --- | --- | --- | --- |
| IFNG | Interferon gamma | Hypermethylated in LTBI | Loss of DNA methylation |
| RASAL1 | RasGAP-activating-like protein 1 | Hypermethylated in LTBI | Loss of DNA methylation |
| GIMAP7 | GTPase IMAP family member 7 | Hypermethylated in LTBI | Loss of DNA methylation |
| ADCY3 | Adenylate cyclase type 3 | Hypermethylated in LTBI | Loss of DNA methylation |
| ATXN1 | Alterernative reading frame 1 | Hypermethylated in LTBI | Loss of DNA methylation |
| DIABLO | Direct IAP-Binding Protein With Low PI | Hypermethylated in LTBI | Loss of DNA methylation |
